# Aerosol Generation from Different Wind Instruments

**DOI:** 10.1101/2020.08.03.20167833

**Authors:** Ruichen He, Linyue Gao, Maximilian Trifonov, Jiarong Hong

**Affiliations:** Department of Mechanical Engineering, University of Minnesota, Minneapolis, MN, USA 55414; Saint Anthony Falls Laboratory, University of Minnesota, Minneapolis, MN, USA 55414

**Keywords:** aerosol transmission, aerosol concentration, aerosol size distribution, wind instruments, articulation, dynamic level

## Abstract

The potential airborne transmission of COVID-19 has raised significant concerns regarding the safety of musical activities involving wind instruments. However, currently, there is a lack of systematic study and quantitative information of the aerosol generation during these instruments, which is crucial for offering risk assessment and the corresponding mitigation strategies for the reopening of these activities. Collaborating with 15 musicians from the Minnesota Orchestra, we conduct a systematic study of the aerosol generation from a large variety of wind instruments under different music dynamic levels and articulation patterns. We find that the aerosol concentration from different brass and woodwinds exhibits two orders of magnitude variation. Accordingly, we categorize the instruments into low (tuba), intermediate (bassoon, piccolo, flute, bass clarinet, French horn, and clarinet) and high risk (trumpet, bass trombone, and oboe) levels based on a comparison of their aerosol generation with those from normal breathing and speaking. In addition, we observe that the aerosol generation can be affected by the changing dynamic level, articulation pattern, the normal respiratory behaviors of individuals, and even the usage of some special techniques during the instrument play. However, such effects vary substantially for different types of instrument, depending on specific breathing techniques as well as the tube structure and inlet design of the instrument. Overall, our findings can bring insights into the risk assessment of airborne decrease transmission and the corresponding mitigation strategies for various musical activities involving wind instrument plays, including orchestras, community and worship bands, music classes, etc.

## 1. Introduction

The coronavirus 2019 (COVID-19) pandemic has caused major disruption to our economy and social activities (Nyenhuis, Greiwe, Zeiger, Nanda, & Cooke, 2020; Seetharaman, 2020), particularly to the entertainment industry, leading to a total shutdown of music production and public events (France, 2020; Garcia et al., 2020). While businesses have started to recover, as of today, with stringent preventive measures in place to minimize risk (Bartoszko, Farooqi, Alhazzani, & Loeb, 2020; Cheng et al., 2020; Sommerstein et al., 2020), many such measures, including mask-wearing, cannot be readily implemented to musical activities. Continuing these activities without appropriate risk assessment and mitigation strategies can result in devastating consequences (Chief, 2020; Lint, 2020). Specifically, a choir from Mount Vernon, Washington, which kept a regular rehearsal for three weeks, had 45 members diagnosed with COVID-19, and two died even with a standard social distancing followed (Chief, 2020). Another choir from Amsterdam experienced a similar tragedy (102 out of 130 members fell ill, and four died) with a late cancellation of their performance and without keeping a suggested social distance (Lint, 2020). Such incidences provide strong evidence on the airborne transmission of COVID-19, which has gained more and more recognition through a series of follow-up studies with controlled experiments (Lednicky et al., 2020; Sia et al., 2020). Particularly, as singing is shown to produce significantly more aerosols than normal speaking and breathing characterized in the literature (Johnson et al., 2011; Loudon & Roberts, 1968; Mürbe, Fleischer, Lange, Rotheudt, & Kriegel, 2020; Sommerstein et al., 2020), the aerosols from an asymptomatic singer can contain viruses, potentially causing a “super spread” of COVID-19 in the proximity of the singer. Similar to singing, it raises serious concerns that an exceedingly high dose of aerosols could be generated by musicians playing various woodwind and brass instruments like that (~10^4^ times of normal breathing) reported for vuvuzela (Lai, Bottomley, & McNerney, 2011). However, so far, very few studies have investigated the aerosol generation and assessed the corresponding risks during these instrument plays. The existing ones related to this topic focus on examining airflows induced by such instruments (Becher, Gena, & Voelker, 2020; Kähler & Hain, 2020; Spahn & Richter, 2020). Becher et al. (2020) and Kähler & Hain (2020) showed that brass instruments only influence flow within 0.5 m from the outlet while woodwind, especially flute, can yield flow movement distance over 1.0 m beyond the outlet. Spahn & Richter (2020) evaluated the risk based on such flow influence zone and provided the spacing rules for safer performance. Collaborating with 15 musicians from the Minnesota Orchestra, our work provides the first systematic examination of aerosol generation from 10 woodwind and brass instruments (Fig. S1) under music with various dynamic levels and articulation patterns (Table 1) as well as the risks associated with these instrument plays. We not only reveal the orders of magnitude variability of aerosol generation across different instruments and musical patterns but also elucidate the connection of such aerosol generation with the instrument design and breathing characteristics under different music plays. Our findings can be further generalized to other woodwind and brass instruments not included in the present study and for the safe arrangement of different musical performance settings.

**Table 1.**
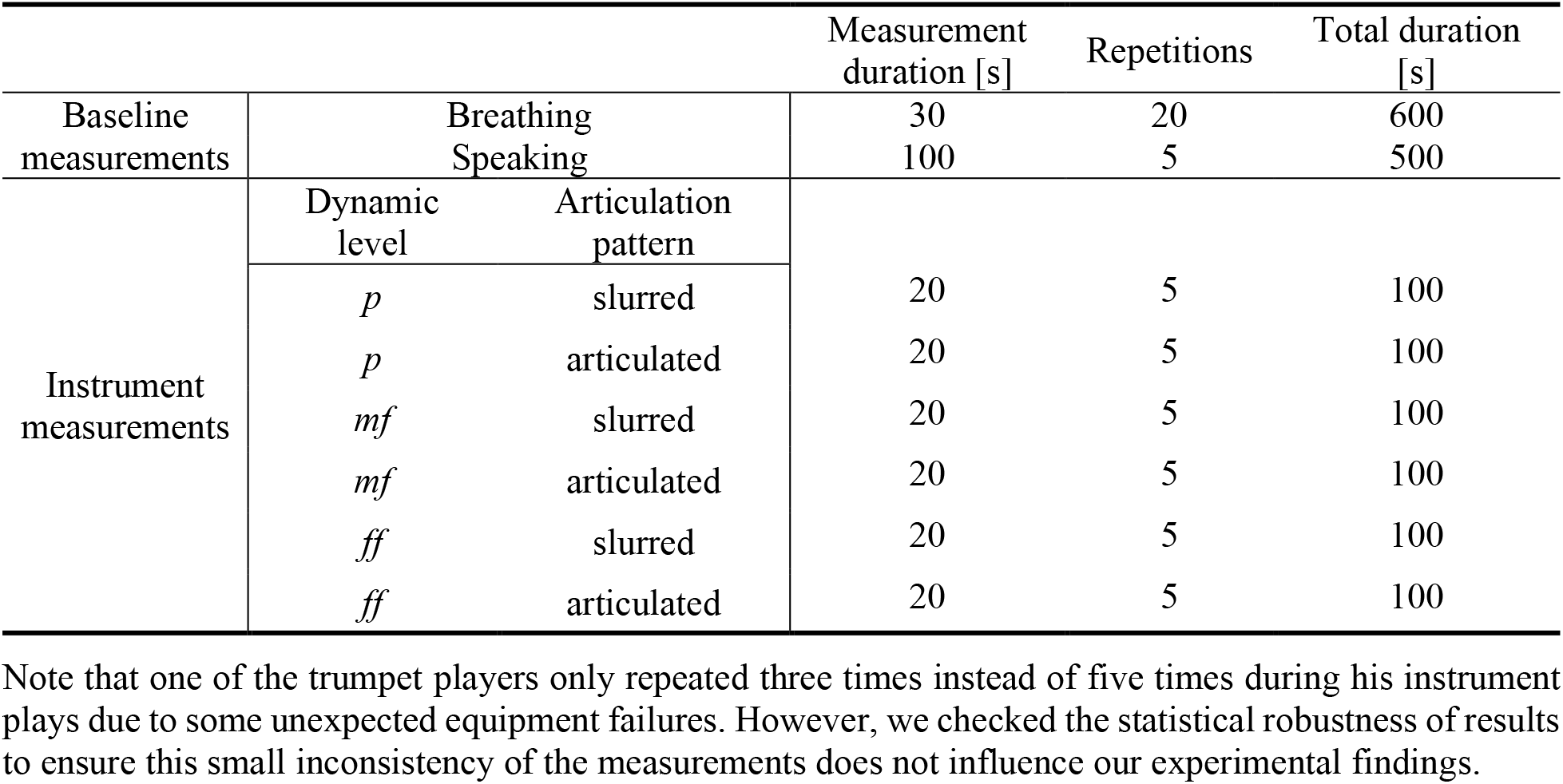
A summary of all the measurement cases for each participant.

## 2. Methods

### 2.1 Participants and involved instruments

Our experiment selects 15 healthy musicians (two females and 13 males) from the Minnesota Orchestra aged between 35 and 60 as participants. Involved musical instruments include trumpet, bass trombone, French horn, tuba, flute, piccolo, bassoon, oboe, clarinet, and bass clarinet, in total 10 types of musical instruments, which cover nearly all types of wind instruments used in Orchestra. Among them, six types of instruments, including trumpet, French horn, flute, bassoon, oboe, and clarinet, are played with two musicians. Each instrument performance consists of six cases with the combination of three dynamic levels and two articulation patterns (Table 1), covering the typical performance range of each instrument. For flute, additional tests are conducted for two special techniques, i.e., tongue ram and jet whistle (with two variations).

### 2.2 Experimental setup

An aerodynamic particle sizer (APS, TSI model 3321) is used to measure the aerosol concentration and size simultaneously under different respiratory activities, i.e., breathing and speaking, which serve as baseline measurements and instrument performance (Table 1). The aerosol size measurement of APS is calibrated to increase the accuracy using digital inline holography (DIH), i.e., a high-resolution imaging system for *in situ* measurements of the particle distribution.

#### APS Measurements

APS measures particle size ranging from 0.5 μm to 20 μm. APS equips an inner pump to provide a sucking-in flow with a flow rate of 5 L/min. Only about 20% of the aerosols remain as sample flow for the measurements, and the rest of them are filtered as sheath flow. The sample flowrate is measured using a Gilibrator (Sensidyne Gilian Gilibrator-2), and it is 1.65 L/min (not 1 L/min mentioned by the manufacture manual). The aerosol concentration is the ratio of a raw count of aerosols (in a unit of particles/L) over the sample flowrate of the APS (in a unit of L/s), according to Eq. (1)

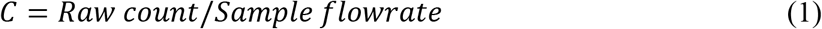

where *C* is the equivalent concentration in a unit of particles/L.

It should be noted that the real emission rate (in a unit of particles/s) has a relationship with the raw count of aerosols following Eq. (2).

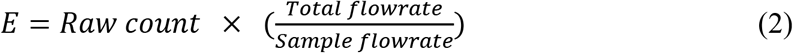

where *E* is the corrected emission rate in the unit of particles/s, and the ratio of total flowrate over sample flowrate is (5/1.65 = 3.03).

As shown in Fig. S2a, a plastic Y-funnel is used to collect particles, which has been used in related aerosol studies (Asadi et al., 2019, 2020). Its convergent channel can improve the uniformity of the airflow, which lowers the chances of aerosol deposition. The funnel size is adjusted accordingly to ensure all particles are collected for each experiment (i.e., breathing, speaking, and playing instruments with varying outlet sizes). Specifically, there are three-size funnels (i.e., small: 102 mm in diameter and 114 mm in height; medium: 140 mm in both diameter and height; large: 190 mm in diameter and 178 mm in height). A small size funnel is used for breathing and speaking experiments. For the instrument experiments, as shown in Fig. S2b, we place the medium and large funnels close to the instrument outlets and cover the outlets without direct contact to minimize the aerosol deposition for most instruments (i.e., trumpet, piccolo, flute, oboe, bassoon, bass clarinet, clarinet). For those instruments with outlets much larger than the funnel (i.e., bass trombone, tuba, French horn), we place the small-size funnel several centimeters inside the instrument outlets (i.e., bells) without direct contact. In addition, to minimize the resistance of the funnel to the flow, the centers of instrument outlets and the funnel are aligned.

A 500-mm long silicon tube connects the funnel with APS for both baseline and instrument measurements. The inner diameter of the tube matches the inlet of the APS of 12 mm. The conductive tube is selected to minimize the aerosol depositions associated with electrostatic charge during the transport, following the APS manufacturer’s instruction (TSI, 2004). After each test, we monitor the concentration status LED signal of the APS to confirm there is no liquid film formation inside the tube. Note that the APS equips an inner pump to suck the aerosols into it with a total air flowrate of 5 L/min, and the corresponding flow speed inside the tube is about 0.74 m/s, which is much higher than the flow speeds measured at outlets of the 10 musical instruments (~ 0.25 m/s on average, measured at the outlet center using an anemometer). The velocity difference provides a negative pressure zone near the funnel stem and tube, which guides the aerosols moving into the APS and further minimizes the influence of aerosol leakage, aerosol deposition in the funnel, and different flow profiles from the instruments on the final measurement results.

To mitigate the measurement uncertainty induced by extra movements during each test, a tripod/hand is used to support the tube and funnel at a certain height. Such height can be adjusted to accommodate different respiratory activities, including breathing, speaking, and playing instruments. All the measurements are conducted in a room of 4.6 m (length) × 3.7 m (width) × 3.4 m (height). The room is maintained with a low environmental dusty level of 0.6 particles/s (significantly lower than the aerosol concentration from breathing, measured by APS of the ambient environment) using a HEPA Air Scrubber (MultiPro Model AH2000). During the tests, the funnel inlet is largely blocked by the participant’s face or the instrument outlet, which further reduces the influence of ambient particles on the APS measurements.

#### APS calibration using DIH

APS measurements tend to provide a smaller size for liquid aerosols due to evaporation (Johnson et al., 2011; Morawska et al., 2009) and have uncertainties in particular size measurements due to the aerosol deformation (Baron, 1986; C. J. Tsai, Chen, Huang, & Chen, 2004) and uncertainties in flowrate quantification (Pfeifer et al., 2016). To eliminate the influence of such uncertainties on the measurements, we employ digital inline holography (DIH) technique, i.e., a high-resolution 3D imaging system, for the calibration of the APS measurements. DIH uses a coherent light source (e.g., laser) to illuminate a sample volume and a camera to capture the patterns generated from interference between the light scattered by objects in the sample and the un-scattered part of the light, referred to as holograms hereafter (Katz & Sheng, 2010). The recorded holograms are then numerically reconstructed using different diffraction kernels to obtain in-focus objects within the sample volume. DIH has been broadly used as a high-fidelity tool for *in situ* measurements of particle concentration, size, and shape over a broad range of applications (Beals et al., 2015; Graham & Nimmo Smith., 2010; Katz & Sheng, 2010; Kumar et al., 2019; Li, Miller, Wang, Koley, & Katz, 2017; Shao, Li, & Hong, 2019; You et al., 2020). Particularly, DIH has been recently used for *in situ* measurements of aerosol generation from human exhalation, capable of capturing the presence of droplets and crystalline particles in the respiratory gas flow as well as their concentration and size distribution (Shao et al., 2020). Specifically, in our experiments, the DIH system (shown in Fig. S2c) consists of a He-Ne laser source (632 nm wavelength), a three-axis spatial filter, a collimation lens, a 10X infinity-corrected objective, and a camera operating with 2000 pixels x 2000 pixels active sensor with 10 frames/s. The calibration is conducted by applying both APS and DIH to measure the aerosols generated by a nebulizer (TSI Single Jet Atomizer 9302) under different back-pressure conditions (Table S1).

The size distribution histograms measured with APS and DIH are fitted using log-normal distributions, and the corresponding mapping in terms of the transformations of aerosol size between the two systems is derived. The calibrated aerosol diameter (*D*_P0_) is obtained with the aerosol diameter measured by APS (*D*_P_) based on Eq. (3).

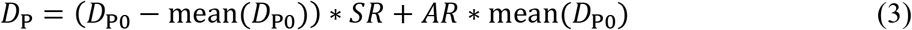

where *AR* and *SR* represent the amplifying ratio and shrink ratio in the calibration (summarized in Table S1), respectively.

### 2.3 Breathing and speaking measurements

The measurements of aerosol generation during normal breathing and speaking of the participants are conducted to serve as baseline measurements to compare with those from the instrument play. In the breathing measurements, the breathing frequency of each participant is synchronized using a metronome at 76 beats. The same breathing pattern of two-beat nose inhalation followed by three-beat mouth exhalation is used for each participant, similar to patterns used in the related studies (Asadi et al., 2019; Shao et al., 2020). The corresponding breathing frequency for this breathing pattern is 15.2 breaths/min breathing frequency, placed within the normal breathing range (Tobin et al., 1983). Each breathing run lasts 30 s and is repeated for 20 times. It should be noted that in between every five runs, the participant takes a one-minute break. During the break time, we check the funnel to ensure that there is no visible vapor condensation on the funnel surface. In the speaking measurements, the participant counts from 1 to 100 continually following a timer placed in front of he/her, which is a common test used in the study of aerosol generation from speaking (Chao et al., 2009; Loudon & Roberts, 1967). Each speaking run lasts 100 s and is repeated five times with one-minute rest in between. During these measurements, the sound level of each participant is recorded using a Decibel meter to help the participant maintain a level near 80 dB. It should be noted that the funnel used in the breathing and speaking measurements is 100 mm in bell diameter and a 75 mm distance between the funnel inlet and outlet. During the tests, the participant places his/her nose above the upper edge of the funnel to ensure that only aerosols generated through the mouth are collected and measured by the APS. In addition, the participant is instructed to avoid contacting the funnel surface during the measurements to prevent vapor condensation caused by the full sealing of the funnel. Furthermore, to reduce the ambient airflow into the device, a face mask is used to cover the bottom half of the funnel, as shown in Fig. S2a.

### 2.4 Instrument measurements

We conduct aerosol measurements at the locations where the dominant (i.e., at least one order of magnitude higher than other locations) aerosol leakage occurs during instrument plays using the same standard music sample for all the instruments. The locations of dominant aerosol leakage are determined through an initial probe of all possible air leakage locations for each instrument, including main air outlets, in the vicinity of keyholes, and near the embouchure hole. For all the brass and reed woodwinds, aerosols are primarily generated from their main outlets. For air-jet instruments (i.e., piccolo and flute), we find that the aerosol concentrations from outlets and near the embouchure hole are comparable. Therefore, the measurements are conducted at both locations to make a detailed assessment of aerosol generation during flute plays.

The music sample, i.e., two-octave B flat major scale, is used by each participant to make a consistent comparison of aerosol generation across different instruments. The sample is played under three dynamic levels (loudness from *p*: soft to *mf*: loud and then to *ff*: very loud) and two articulation patterns (slurred - connected and smooth, and articulated- short and separated), a combination of which constitute six test cases including *p*-slurred, *p*-articulated, *mf*-slurred, *mf*-articulated, *ff*-slurred, and *ff*-articulated, as summarized in Table 1. Note that no accented notes are used in articulated plays. During the tests, to keep the consistency among all the participants, the participant plays the instrument following a metronome with two notes per beat. After 8 s instrument performance (16 notes), the participant is instructed to hold the instrument at the same location without blowing for another 12 s to ensure that APS collects all aerosols. Each test case is repeated five times. In the instrument tests, funnels with different sizes are used according to the instrument outlet dimension to ensure that the aerosols emitting from the outlets are completely collected.

In addition, following the suggestion of musicians, we have conducted aerosol measurements near the keyholes during bassoon plays with two special music pieces that are likely to cause more air leakages near the lower keyholes due to frequent usage of corresponding keys. The pieces selected are the first 17 s of the Bassoon Concerto in B-Flat Major (W.A. Mozart), K. 191, Allegro, and the first 13 s of the bassoon cadenza in the second movement of Scheherazade, i.e., Kalendar Prince.

## 3. Results

### 3.1 Influence of musical instrument type on aerosol generation

The aerosol concentration generated from 10 orchestra instruments exhibits two orders of magnitude variation (Fig. 1a), i.e., from ~20 to ~2400 particles/L covering the range of normal breathing (~90 ± 65 particles/L) and speaking (~ 230 ± 95 particles/L) in the present study. Specifically, tuba produces fewer aerosols than normal breathing, while the concentrations from piccolo, flute, bass clarinet, French horn, and clarinet stay within the range of normal breathing and speaking. The musicians playing trumpet, oboe, and bass trombone tend to generate more aerosols than speaking. Accordingly, as higher aerosol concentration leads to increased risk of airborne disease transmission (Fennelly, 2020), we categorize these instruments into low, intermediate, and high-risk levels based on a comparison of their aerosol concentrations with the concentration span of normal breathing and speaking. Note that only the aerosols measured at the main outlet of each instrument are used for comparison in this figure for consistency. For the air-jet instruments (flute and piccolo), the aerosol leakage near the embouchure hole contributes to about half of the total aerosol generation, as shown in Fig. S4. For bassoon plays using special pieces that involve frequent usage of lower keys, the leakage near the corresponding keyholes is also significant and contributes to about 40% of the total in the test cases (Fig. S4). Nevertheless, with the consideration of the abovementioned special cases, the risk categorization of these instruments remains the same. Furthermore, we attribute the large variability of aerosol concentration from different instruments to the combined effects of sound production mechanisms associated with instrument type (i.e., brass and woodwind), mouthpiece (i.e., air-jet, single reed, and double reeds, Fig. S5), and tube structure (i.e., tube length, turnings, and valves) of each instrument. The former two factors affect how aerosols are injected into the instrument, while the latter influences aerosol transport inside the instrument tubes. Specifically, for brass, we observe that the rank of aerosol concentration, i.e., trumpet > bass trombone > French horn > tuba, is inversely correlated with the total tube length of each instrument (Table S2). As aerosols transport in the tubes, they can deposit at their inner surfaces. A longer and narrower tube yields a higher probability of such deposition, leading to a lower aerosol concentration at the outlet (Fennelly, 2020). For woodwinds, we find that their aerosol concentration seems to be strongly influenced by its mouthpiece design. In particular, the air-jet woodwinds (i.e., piccolo and flute) produce the lowest concentration as the respiratory jet impinges the edge of the embouchure hole at a steep angle leading to high particle deposition near the inlet (Fig. S5a), similar to the aerosol deposition (0.1-100 µm with a flow rate of 10-100 L/min) in a cascade impactor (P. J. Tsai, Uang, Wang, Wu, & Shih, 2012). Such deposition due to direct impingement reduces gradually going from air-jet to single-reed (i.e., clarinet and bass clarinet), then to double-reed woodwinds (i.e., oboe and bassoon) as the flow into the main tube becomes more and more aligned the tube axis (Figs. S5b-c). Such process is analogous to spray deposition, where deposition efficiency decreases as the impingement angle of the spray lowers (Gilmore, Dykhuizen, Neiser, Roemer, & Smith, 1999). Moreover, different from other instruments, air-jet woodwinds have air leakage near the mouths during the regular plays (Fletcher & Rossing, 1991), which further reduces the concentration of aerosols from the instrument outlet (Fig. S5a). Besides the mouthpiece, tube structure also plays a role in the aerosol generation of woodwinds. Particularly, bassoon, despite being a double-reed, exhibits the lowest concentration among the six woodwinds, mainly associated with its significantly longer tube. Similarly, for the two single-reed woodwinds, bass clarinet produces fewer aerosols than clarinet due to its extended tube length. For air-jet woodwinds, although the tube length of piccolo is only half of flute, its bore size is half of flute, therefore, leading to a comparable number of aerosols deposited in the tube as flute. Finally, comparing the brass and woodwind with less than 7% difference in tube length, we find that bass trombone has an order of magnitude higher aerosol concentration than bassoon. Such observation can be explained by the specific mouthpiece (i.e., circular openings to the main tubes via a semi-spherical or conical cavity (Fletcher & Rossing, 1991) of brass that results in considerably less deposition as the aerosols enter the tube in comparison to reed instruments (Fig. S6).

**Fig.1.**
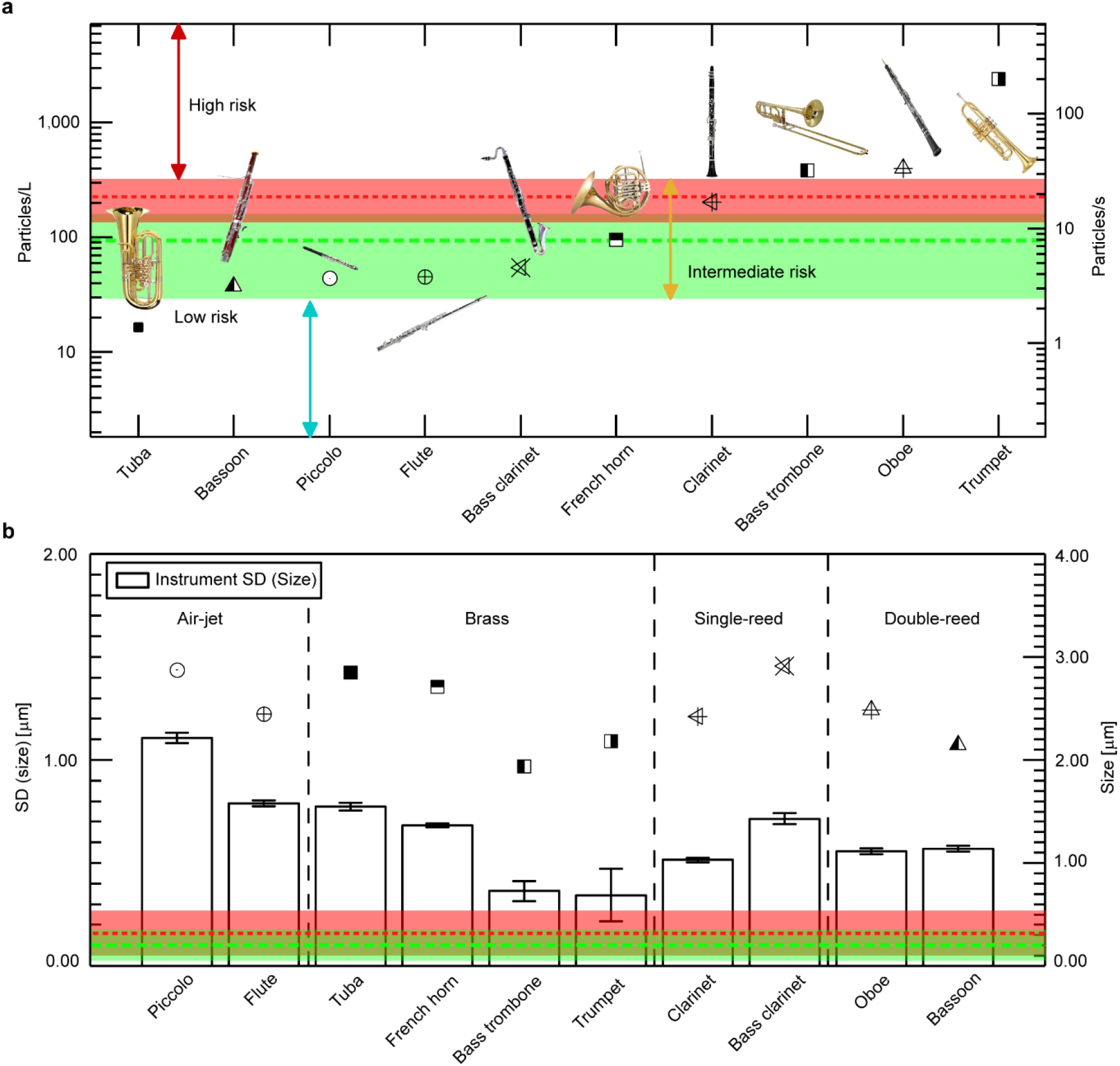
The concentration and size distribution of aerosols generated from different music instrument plays. (**a**) Aerosol concentration levels measured at the outlets of 10 orchestra instruments. Such levels are averaged across different dynamical levels, articulation patterns, and individuals. The green and red dashed lines mark the concentrations of breathing and speaking averaged over all participants, respectively (see Fig. S3 for the breathing and speaking results for musicians playing each instrument). The corresponding shaded regions represent the standard deviation (SD) of these measurements. (**b**) The average size (symbols) and size standard deviation (columns) of the aerosols generated during different instrument plays in comparison to and the corresponding breathing and speaking results (i.e., average and standard deviation illustrated in the same fashion as those in Fig. 1a). The error bar for the column is ± 1 standard deviation obtained using a bootstrap analysis by randomly selecting 80% data to represent the uncertainty in ensemble-averaged statistics of the size variation here. Note that only the aerosols measured at the main outlet of each instrument are used for comparison in this figure for consistency.

In contrast to concentration, the size distributions of aerosols from all the brass and woodwinds are all approximately log-normal (Fig. S7), and their averages are within a close range, i.e., 1.9-3.1 μm (Fig. 1b). Remarkably, the span of aerosol size (characterized using the size standard deviation) during instrument plays are generally broader than those from normal breathing and speaking. We attribute such trend to the more vigorous exhalation involved in instrument plays (Macfie, 1966), which can lead to the increase in size variation similar to those observed in heavier breathing (Asadi et al., 2019). In addition, we find that the span of aerosol size varies more significantly across different instruments compared to their average sizes. Particularly, for air-jet woodwinds of similar straight tube structure and producing aerosols of similar concentration, the span of aerosol size from flute is smaller than piccolo (Fig. S7i, j) owing to its longer and wider tubes. Under such circumstances, aerosols transported in flute have longer residence time, and aerosols of larger size are more likely to deposit on the tube inner surfaces (Table S3). However, this size-dependent deposition does not seem to explain the trends observed in the other instruments with more complex tube and inlet structures, i.e., reeds, turnings, and valves. Instead, we observe a clear negative correlation between aerosol concentration and size variation for instruments of similar types (i.e., brass and reed instruments) due to the span increase resulted from a reduction in sample size (Watkins, 2017). This result suggests that with the increasing complexity of instruments, the deposition appears to be insensitive to the size within the range of our measurements (i.e., 1.4 to 20 μm), as reported (Asmatulu et al., 2017; Heyder, Gebhart, Rudolf, Schiller, & Stahlhofen, 1986).

### 3.2 Influence of dynamic level and articulation pattern on aerosol generation

We investigate the aerosol production when playing different music using six music pieces generated from a combination of three dynamic levels (*p*: soft, *mf*: medium loud, and *ff*: very loud) and two articulation patterns (slurred and articulated). We find that the influence of dynamic levels varies according to different instrument types (Fig. 2a). Based on the correlation of aerosol concentration change with increasing dynamic level, we categorize the instruments into four groups, i.e., positive correlation (denoted as *p* < *mf* < *ff* including oboe, bassoon, and clarinet), negative correlation (denoted as *p* > *mf* ≳ *ff* including flute and piccolo), and no clear correlation (including *p* < *ff* < *mf* for bass clarinet, trumpet, bass trombone, and French horn, and *p* ≅ *mf* ≅ *ff* for tuba). Such trend can be explained by two competing factors that influence the change of aerosol concentration with an increasing dynamic level. On the one hand, the augment of flow rate associated with increasing blowing pressure at higher dynamic levels (Fletcher & Tarnopolsky, 1999; Fuks & Sundberg, 1996) can lead to more aerosol production, similar to those from heavy breathing (Asadi et al., 2019). On the other hand, higher blowing pressure can also increase the deposition and/or leakage of aerosols due to enhanced flow impingement at the inlets(Gilmore et al., 1999) and inner walls of bifurcating tubes (Sippola & Nazaroff, 2005), particularly for the instruments with convoluted tube and inlet structures, causing a decrease in the aerosol concentration at the outlets. Specifically, for instruments consisting of primarily straight tubes (i.e., clarinet, oboe, and bassoon, see Figs. S1g, i, and j), the increase of aerosol production with dynamic level is dominant, which yields a positive correlation. For air-jet woodwinds, considering the steep-angle impingements at the inlet, the latter factor overwhelms the former, contributing to a negative correlation. For the instruments with more complex tube structures (i.e., brass and bass clarinet, see Figs. S1a-d and h), the competition of both factors complicates the correlation. We observe that nearly no correlation with dynamic level for tuba potentially due to its exclusively long tube and lowest aerosol production, and the peak of aerosol concentration at an intermediate dynamic level for the other instruments in this group.

**Fig. 2.**
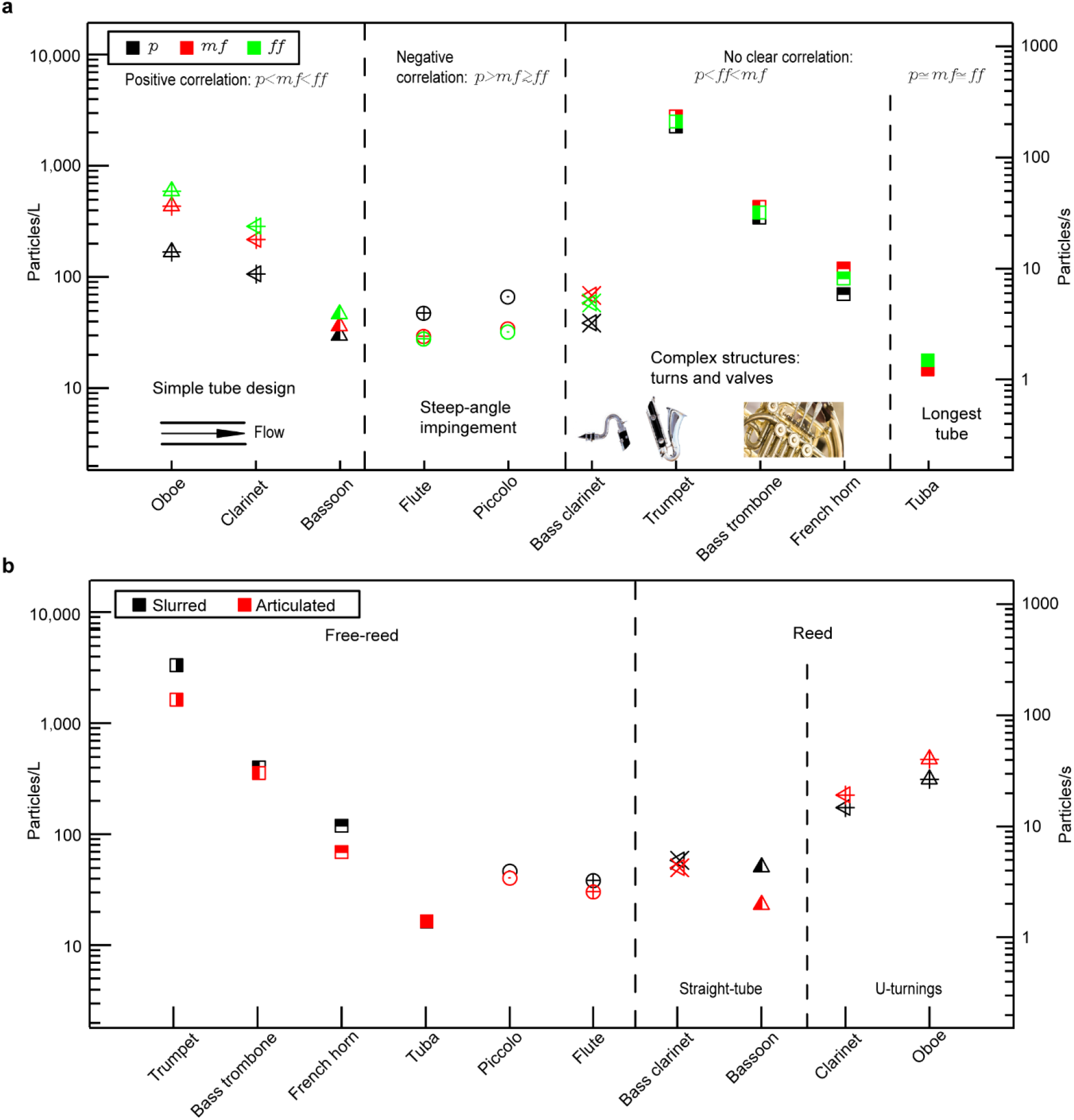
Effects of dynamic level and articulation pattern on the aerosol concentration from different wind instruments. (**a)** Aerosol concentration under three dynamic levels, i.e., *p* (piano, soft), *mf* (mezzo forte, medium loud) and *ff* (fortissimo, very loud), and (**b)** under two articulation patterns, i.e., slurred (sustained pieces) and articulated (separated pieces). In (**a)**, all the instruments are divided into three groups, i.e., positive correlation with dynamic level (denoted as *p*<*mf*<*ff*), negative correlation (*p*>*mf≳ff*), and no clear correlation (including *p*<*ff*<*mf* and *p*≅*ff*≅*mf*), depending on how their aerosol concentrations vary with respect to increasing dynamic levels. In (**b)**, the influence of articulation patterns on aerosol concentration is different for free-reed and reed instruments.

We also examine the variation of aerosol concentration with different articulation patterns, i.e., slurred (connected and smooth notes) and articulated (separate and short notes). For free-reed instruments, the instrument plays with slurred patterns generally lead to higher aerosol production than with articulated at the same dynamic level since slurred notes usually yield a higher average flow rate than the articulated ones at the same dynamic level (Pàmies-Vilà, Hofmann, & Chatziioannou, 2018). Noteworthily, we find that the aerosol concentration from tuba is insensitive to the change of articulation patterns like that for different dynamic levels, potentially due to the same reason. In comparison to free-reed instruments, the influence of articulation is more complicated for reeds. Specifically, although a similar trend (i.e., higher aerosol concentration with slurred pattern) is observed for bass clarinet and bassoon, the concentrations for oboe and clarinet show an opposite dependence on articulation pattern. In general, during the play of reed instruments, the tongue is pressed against reeds to make articulated notes. Such tongue placement is likely to yield higher aerosol production like the higher concentration observed in the speaking of ‘i’ in comparison to ‘a’ (Asadi et al., 2020). However, as aforementioned, the tube and inlet structures can also influence the deposition of aerosols during their transport and, ultimately, the concentration at the outlets. For oboe and clarinet with straight tubes (Fig. S8a), we expect that the tongue movement induced enhancement of aerosol production overwhelms the loss of aerosols during the transport, causing the reversed trend in comparison to that of free-reed instruments. Comparatively, bass clarinet and bassoon with steep turnings (Fig. S8b) may result in stronger aerosol deposition loss, particularly under the intense pressure oscillation during the plays using articulated notes (Pàmies-Vilà et al., 2018). Such loss may offset the aerosol production enhancement induced by tongue movement, lowering the corresponding aerosol concentration at the outlets.

### 3.3 Influence of individual on aerosol generation

We investigate how the aerosol generation during instrument plays is influenced by normal respiratory behaviors, i.e., breathing and speaking, of an individual. Such investigation is conducted for six types of instruments, each of which is played by two musicians (Fig. 3a). For free-reed wind instruments (brass and air-jet woodwinds), we find that a positive correlation of the concentration of aerosols generated during instrument play with those from an individual’s breathing and speaking. Such observation highlights that the individuals who naturally produce more aerosols (Asadi et al., 2020), especially those referred to as “super emitters” (Fennelly, 2020), tend to generate more when they play these instruments. However, for reed instruments, the relation between individual respiratory behavior and instrument play becomes more complicated. Specifically, clarinet, oboe, and bassoon yield positive, negative and no appreciable correlations, respectively. We attribute this complication to the specific breathing control used by musicians when playing reed instruments. In particular, different musicians have different performance styles, and they can use lip pressure to control the opening between reed and embouchure edge, instead of controlling the blowing pressure to for the adjustments of dynamic levels or articulation patterns (Fletcher & Rossing, 1991). Such exhalation behavior is significantly different from normal breathing, reducing the influence of individual natural breathing characteristics on instrument aerosol production (Asadi et al., 2020). In addition, the aerosol size variation shows a trend consistent with the concentration, i.e., positive correlation with individual respiratory behaviors for free-reed instruments and no clear correlation for reeds (Fig. 3b), which consolidates the abovementioned explanation. Specifically, for the free-reed instruments, the musicians that tend to generate aerosols in a broader range of sizes also present larger aerosol size variation in their instrument plays. In contrast, for the reed instruments that require the special lip pressure control during the play, no consistent correlation is observed among the three reed instruments (i.e., clarinet, oboe, and bassoon).

**Fig. 3.**
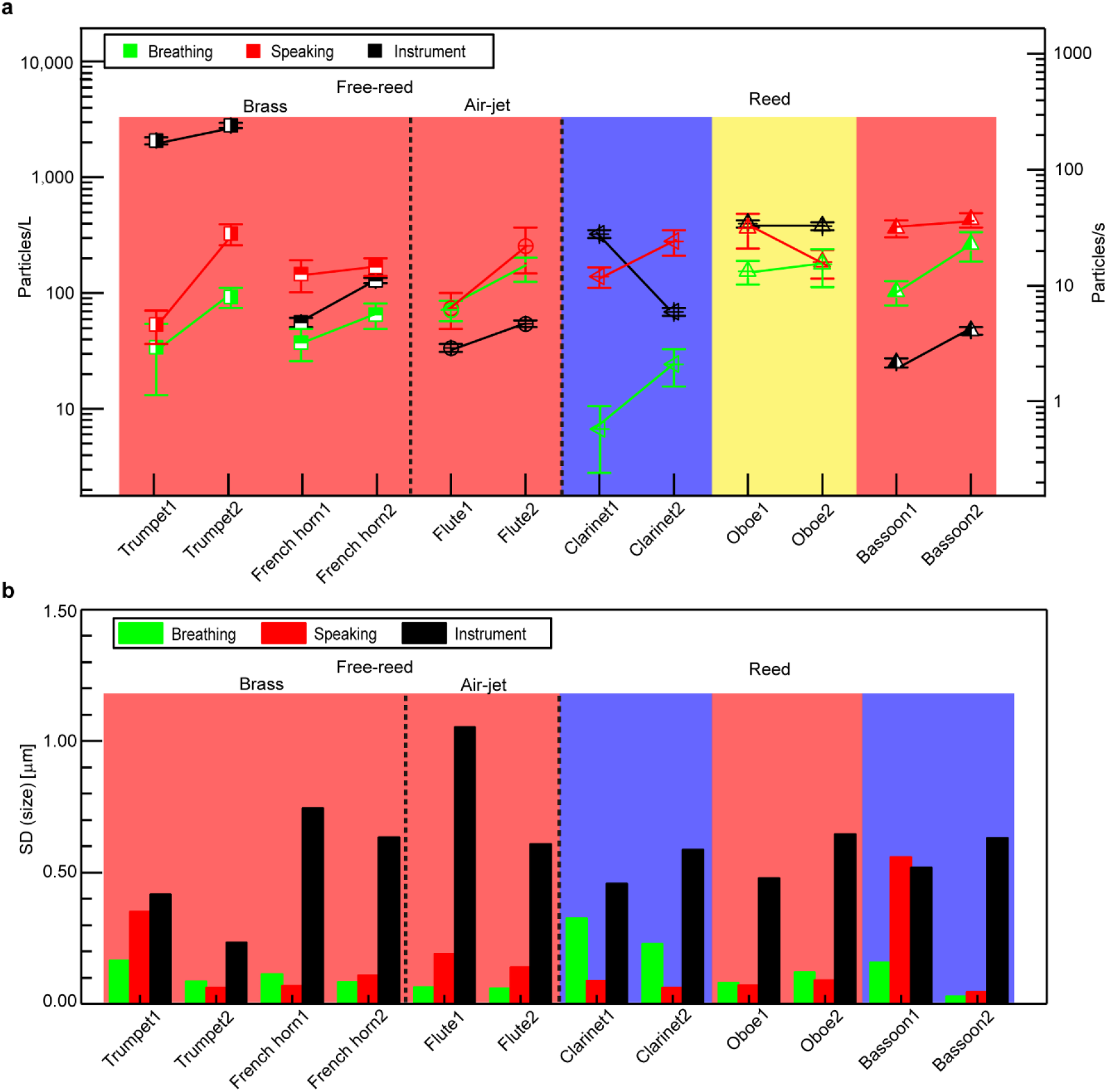
Influence of individual on the aerosol concentration and size variation from different instruments. (**a)** Comparison of aerosol concentrations generated from two musicians playing the same instrument along with their individual breathing and speaking measurements. The red, blue, and yellow shaded areas highlight the instrument plays that show positive, negative, and no correlation with their breathing and speaking behaviors in terms of aerosol generation, respectively. **(b)** Comparison of size variation of the aerosols generated from two musicians playing the same instrument along with their individual breathing and speaking measurements. Note that for French horn, aerosol size variations of breathing and speaking for two participants show diverge trends. We consider the breathing case is more representative than speaking and used for the analysis of individual effect.

### 3.4 Influence of play techniques on aerosol generation

We also examine the influence of special techniques used in flute performance, including tongue ram and jet whistle with two variations, on aerosol generation. Remarkably, in comparison to the aerosol generation using basic flute techniques, we notice a drastic increase of aerosol concentration when using special techniques, i.e., nearly 50 times increase using tongue ram and about five and three times increases using different variations of the jet whistle, respectively (Fig. 4a). Such drastic increases in aerosol generation are inherently associated with exhalation behaviors used for special techniques. These two special techniques require the musicians to completely seal the embouchure hole with lips (Andrus & Shanley, 1980; Heiss, 1972). Such behavior can minimize the air leakage near the mouth and shift the flow impingement point from the edge of the embouchure hole to the inner surface of the main tube, reducing aerosol loss at the inlet. Particularly, for tongue ram, the powerful and rapid tongue propelling into the embouchure hole (Andrus & Shanley, 1980) can further increase the aerosol generation (Bake, Larsson, Ljungkvist, Ljungström, & Olin, 2019). In addition to concentration, the average size and size variation during the flute performance using special techniques decrease compared to those using basic techniques (Fig. 4b). Specifically, the variation of aerosols generated using tongue ram decreases with almost doubled probabilities at a small size range of 1.4 -2.0 μm than the basic performance. Similar trends of increments in the concentration of small size range are also observed in the performance of jet whistles with two variations. The significantly higher concentration and small size of the aerosols when using the special techniques pose higher risks of airborne disease transmission.

**Fig. 4.**
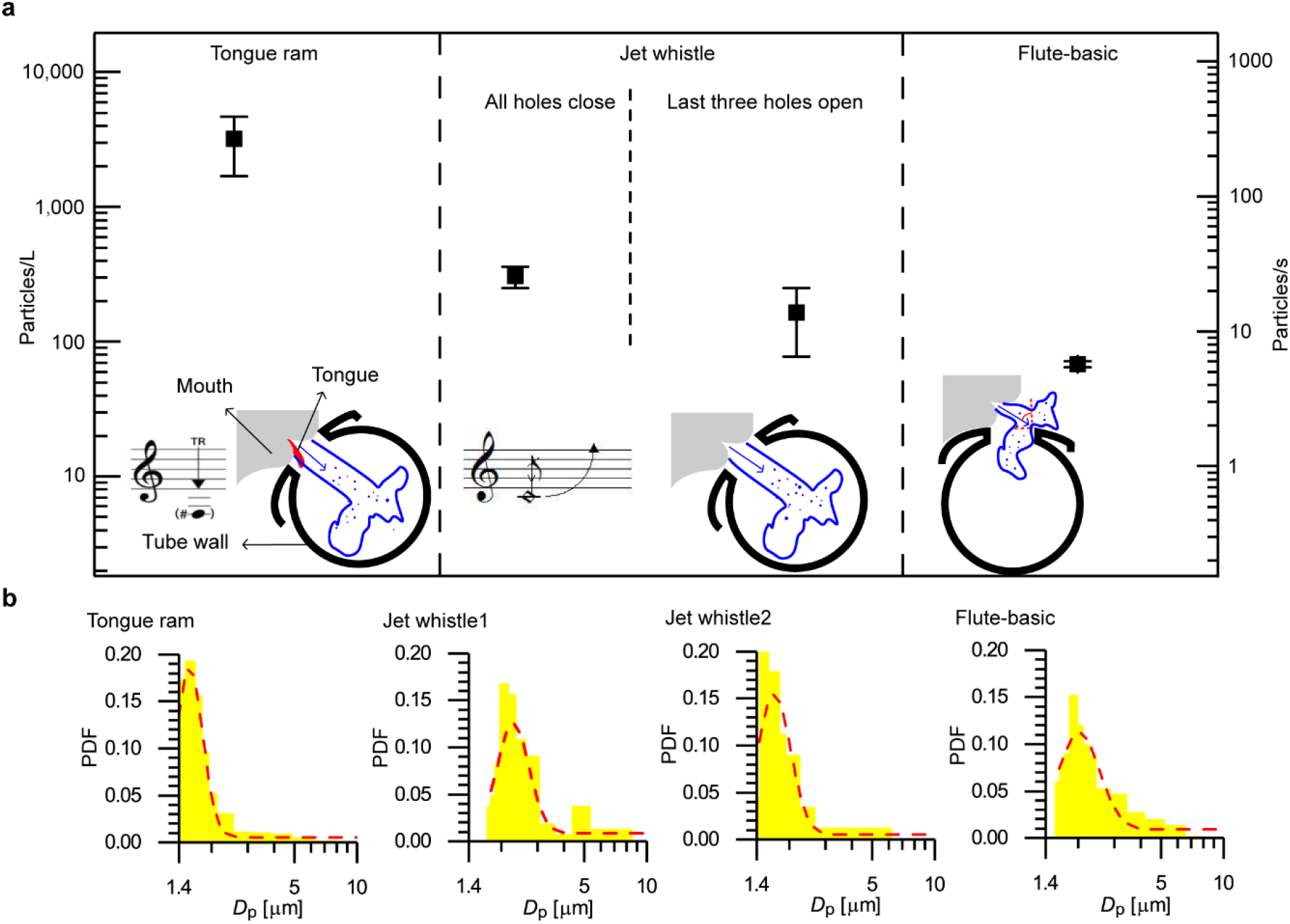
Influence of special techniques on the aerosol concentration for flute performance. **(a)** Aerosol concentrations generated from flute performance using special techniques including tongue ram and jet whistle with two variations (i.e., jet whistle 1 that has all holes closed, and jet whistle 2 that leaves the last three holes open) in comparison to that from the basic technique. Error bars represent the standard deviation of the measurement. The inset figures for each special technique include a schematic illustrating the flow of aerosols into the main tube of the flute and the corresponding music note. **(b)** Probability density functions (PDFs) of the aerosol size from flute performance using basic and different special techniques.

The red dashed line in each histogram is the log-normal fitting curve of the PDF. In total 16 bins are used in the range from 1.4 to 20 μm. Note that the lower bound of 0.5 μm in the raw APS measurement is calibrated to 1.4 μm here based on Eq. (3).

## 4. Conclusions and Discussion

In this study, we provide the first systematic investigation of aerosol generation for a variety of wind instruments through the collaboration of 15 musicians from the Minnesota Orchestra. We find that aerosol generation from wind instrument plays is influenced by a combination of breathing techniques as well as the tube structure and inlet design of the instrument. Our results show that the aerosol concentration from different instrument plays exhibits two orders of magnitude variation. Specifically, tuba produces fewer aerosols than normal breathing, while the concentrations from bassoon, piccolo, flute, bass clarinet, French horn, and clarinet stay within the range of normal breathing and speaking. Trumpet, oboe, and bass trombone tend to generate more aerosols than speaking. Accordingly, we categorize these instruments into low, intermediate, and high-risk levels based on a comparison of their aerosol concentrations with the concentration span of normal breathing and speaking. In contrast to concentration, the size distribution of aerosols from all the brass and woodwinds are all approximately log-normal, and their averages are within a close range, i.e., 1.9-3.1 μm. In addition, we find that the span of aerosol size varies more significantly across different instruments compared to their average sizes. Moreover, the dependence of aerosol production upon the dynamic level and articulation pattern varies for different wind instruments. Particularly, only the instruments with straight tube design show an increase of aerosol concentration with increasing dynamic level, while such trend is reversed for air-jet instruments and becomes unclear for instruments with complex/very long tube structures. For free-reed instruments, the slurred play tends to produce more aerosols than playing articulated notes, while no clear trend with articulation pattern is observed for reeds. Furthermore, we find that the individual’s natural respiratory behaviors can positively influence the aerosol generation during instrument plays. However, such dependence only holds for free-reed instruments, not for reeds due to the special lip control used in the play of such instruments. Finally, we notice a drastic increase in aerosol concentration during flute performance with special techniques, i.e., nearly 50 times increase using tongue ram and about five and three times increases using different variations of the jet whistle, respectively.

The findings in our study can be generalized for understanding and estimating the aerosol generation from other musical instruments that are not included in the present study. For example, single-reed instrument like saxophone with steep turnings near its inlet and outlet is likely to produce a relatively lower level of aerosols, and its aerosol generation may be insensitive to the variation of dynamic level and individual respiratory behaviors like those observed in bass clarinet. The brass instrument like cornet with comparable tube length with trumpet is likely to produce a high level of aerosols, and the performance with cornet may generate more aerosols with slurred notes than using articulated notes and have a positive correlation in individual respiratory behaviors. In general, our findings can provide valuable insights into the risk assessment of airborne disease transmission and the corresponding mitigation strategies (e.g., seating arrangement and ventilation) for different musical activities involving the usage of wind instruments, including orchestras, community and worship bands, classes in music conservatories, etc. Specifically, for the activities involving high-risk level wind instruments (i.e., trumpet, bass trombone, oboe, etc.), extra preventive measures such as a reduction in occupancy, additional social distancing, and ventilation enhancement are needed in comparison to playing instruments with lower risk levels.

Note that different musicians have different performance styles when they are playing the same music piece, particularly for reed instrument plays, which may affect the aerosol generation levels. Specifically, based on the performance styles, musicians have their own preferences on reed selection (e.g., reed width, reed hardness, etc.) that can potentially influence aerosol generation. For example, as to the clarinet performance, some musicians prefer to adjust the blowing pressure to change the dynamic level and articulation pattern, while some alter the mouthpiece pressure instead, which may lead to substantial difference in aerosol generation as shown in the complicated individual effect in Fig. 3. In addition, the hardness of a reed varies as it ages and with the change of temperature and humidity, which may also contribute to variations in aerosol generation. All the aforementioned factors can add uncertainties to our findings. To obtain a more accurate assessment of airborne transmission during wind instrument plays, it is desirable to integrate our findings with the information of instrument-generated flows and ambient flows (including both ventilation and natural convection) under specific settings. Such information can be derived from *in situ* measurements of flow and aerosol transport and the numerical simulations in the future.

## Data Availability

The data will be available upon request.

## Acknowledgments

This work was support by the School of Medicine of the University of Minnesota. The authors would like to thank Dr. Siyao Shao, Santosh Kumar, Buyu Guo and Dr. Kevin Mallery for their assistance in the preparation of the experiments. We would also like to thank Dr. David Y.H. Pui for equipment support, and his postdocs Dr. Qisheng Ou and Dr. Seong Chan Kim and student Dongbin Kwak for their assistance in the equipment calibration. In addition, we would like to thank Mele Willis from the Minnesota Orchestra for arranging the experiments and other 15 musicians for their participation in the experiments. Finally, we would like to thank Mele Willis again and Dr. Robert Patterson for their constructive comments on our analysis.

## Supplementary Information

### Supplementary Figures

**Fig. S1.**
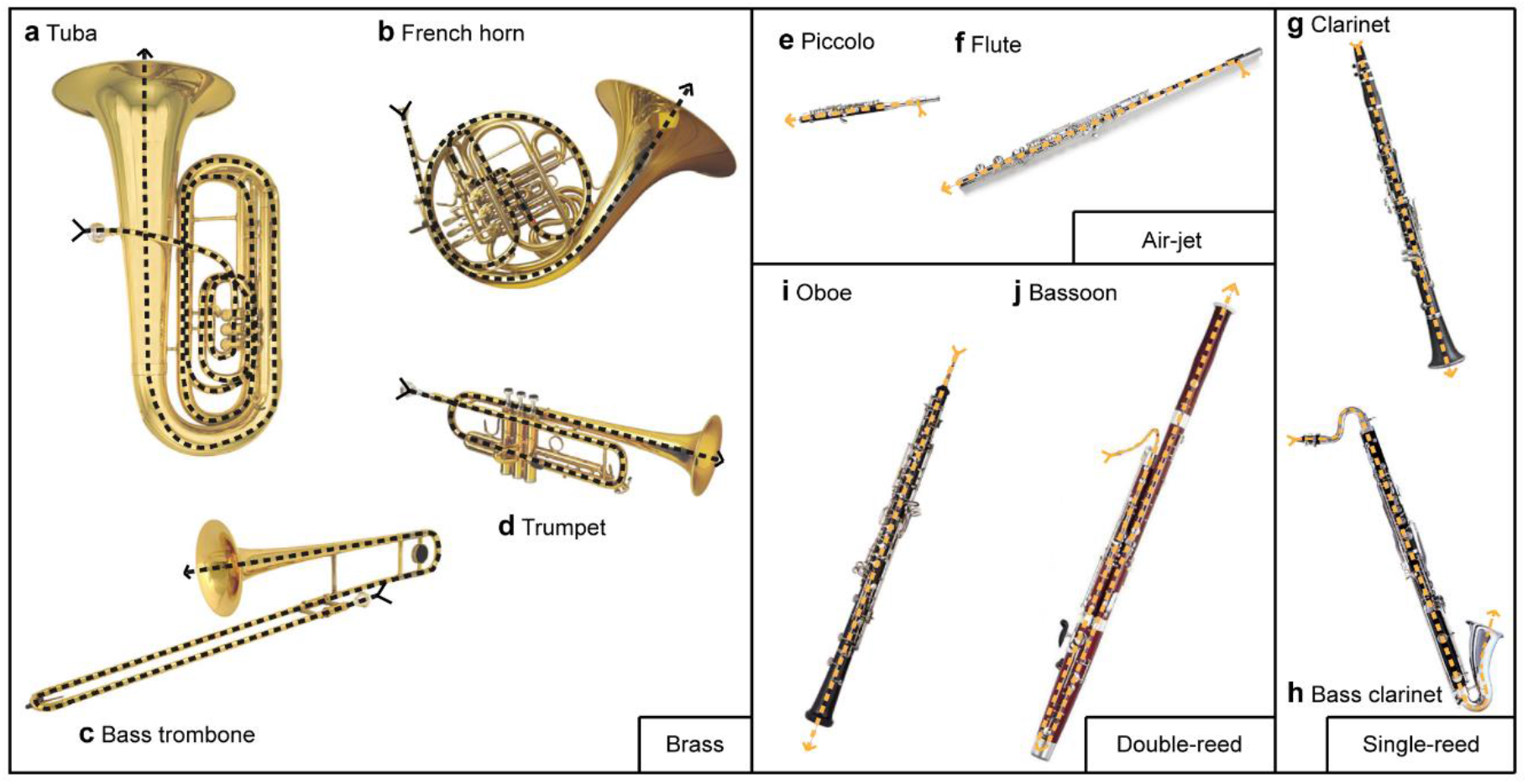
Images of 10 instruments used for the aerosol measurements. Brass instruments include (**a)** tuba, (**b)** French horn, (**c)** bass trombone, and (**d)** trumpet. Air-jet woodwinds include (**e)** piccolo and (**f)** flute. Single-reed woodwinds include (**g)** clarinet and (**h)** bass clarinet. Double-reed woodwinds include (**i)** oboe and (**j)** bassoon. The dashed line marks the main flow path in each instrument.

**Fig. S2.**
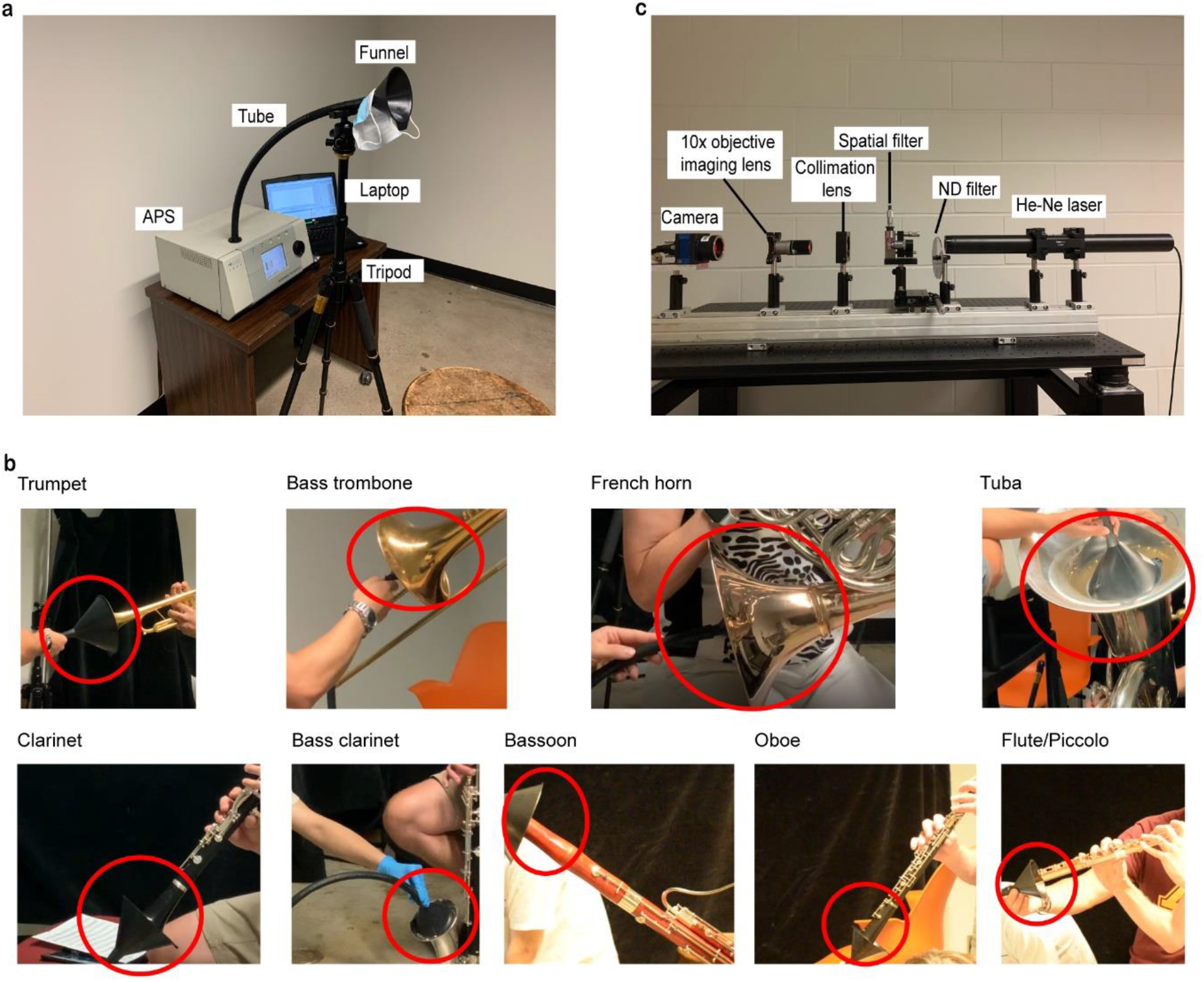
Experimental setups of the Aerodynamic Particle Sizer (APS) measurements and digital inline holography (DIH) system used to calibrate APS. **(a)** APS setup. The funnel is adjusted to fit the different size of orchestra instrument outlets for the instrument measurements. A laptop is used for data acquisition, and a tripod is employed for holding the tube and funnel at the fixed location during the tests. **(b)** Photos of the funnel and instrument outlet for each instrument. The location of funnel is highlighted with a red circle in each photo. **(c)** DIH setup for APS calibration.

**Fig. S3.**
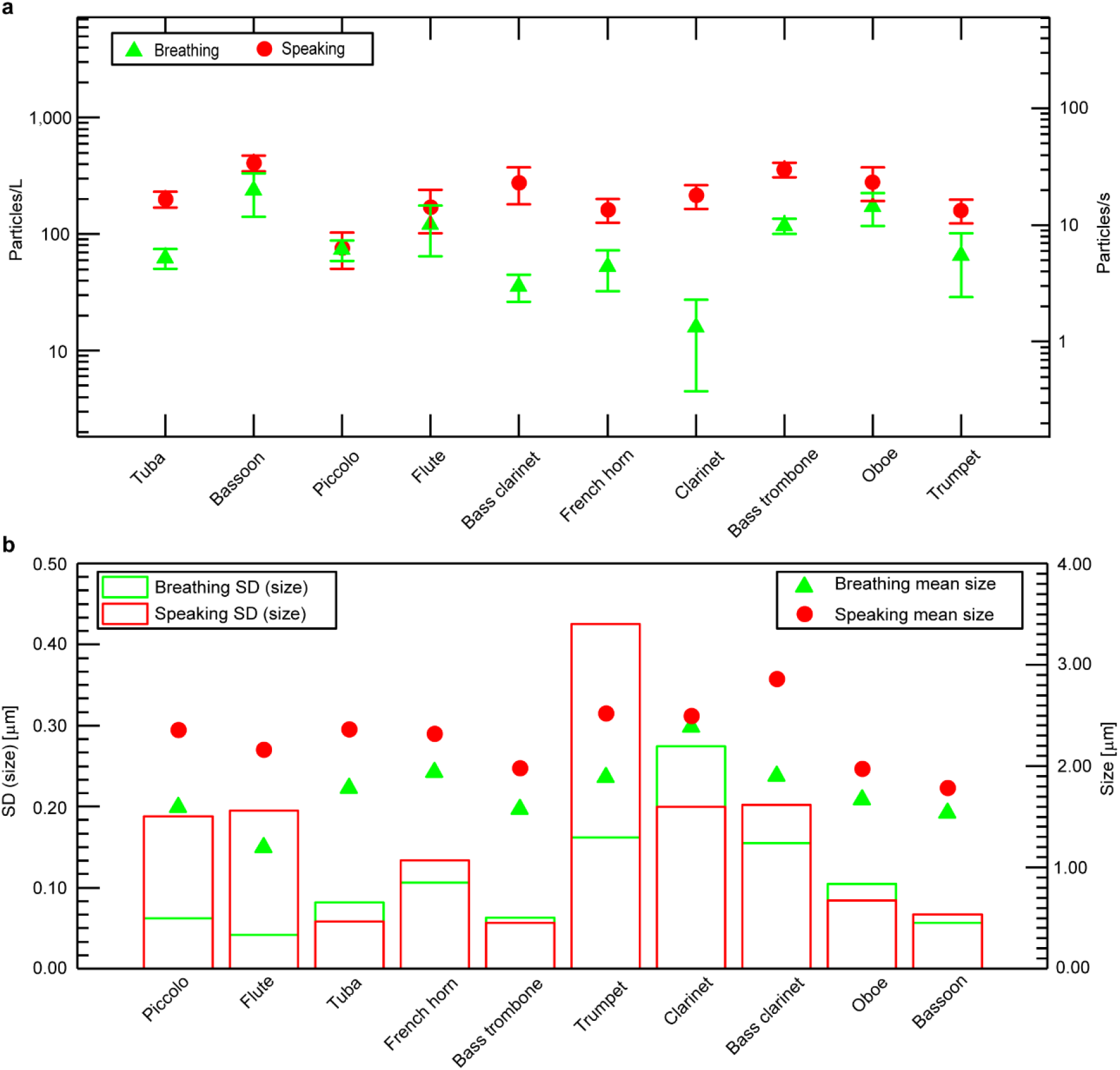
Aerosol concentrations and size of breathing and speaking cases of the participants for each instrument. **(a)** Aerosol concentrations are ranking from lowest to highest. The error bar corresponds to ±1 standard deviation of the dataset. (**b)** The average size and size variation of aerosols of the breathing and speaking tests for 10 instrument cases. The size variation here uses the standard deviation of the dataset.

**Fig. S4.**
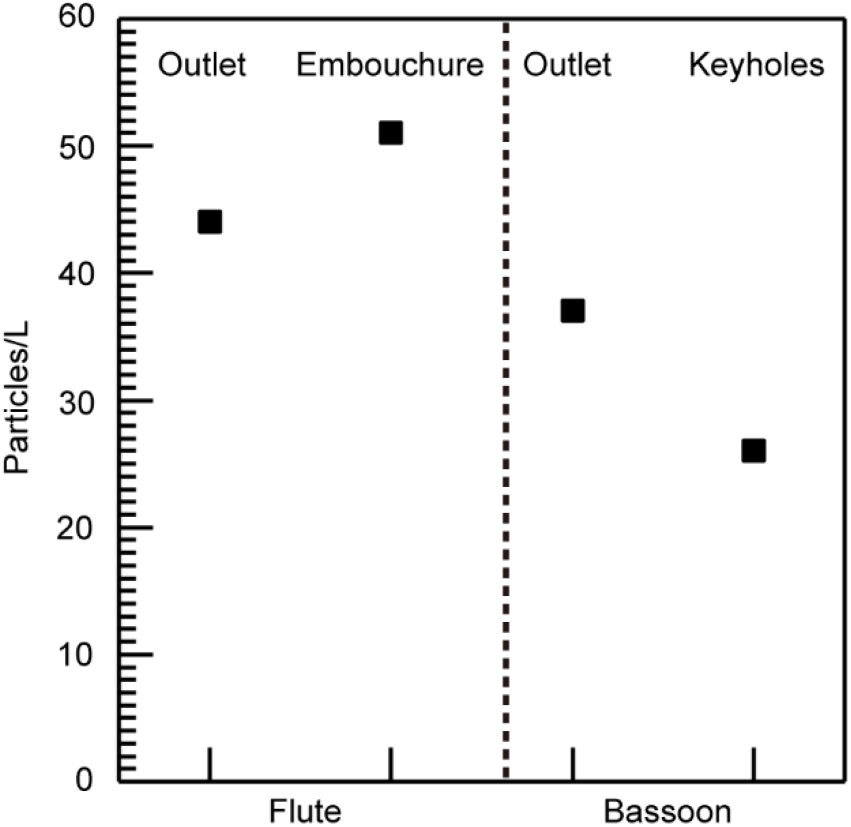
The aerosol concentration measured at different locations during flute and bassoon plays. For flute play using the standard music sample, the aerosol concentrations measured near the embouchure hole and main outlet are 51 particles/L and 44 particles/L, respectively. For bassoon play with special music pieces, the aerosol concentrations measured near the keyholes and main outlet are 26 particles/L (24 particles/L for piece one, and 28 particles/L for piece two) and 37 particles/L, respectively. Note that for the aerosol measurements near the lower keyholes during bassoon plays, two special music pieces which are likely to cause more air leakages near the lower keyholes are selected due to frequent usage of corresponding keys. The pieces used are the first 17 s of the Bassoon Concerto in B-Flat Major (W.A. Mozart), K. 191, Allegro, and the first 13 s of the bassoon cadenza in the second movement of Scheherazade, i.e., Kalendar Prince.

**Fig. S5.**
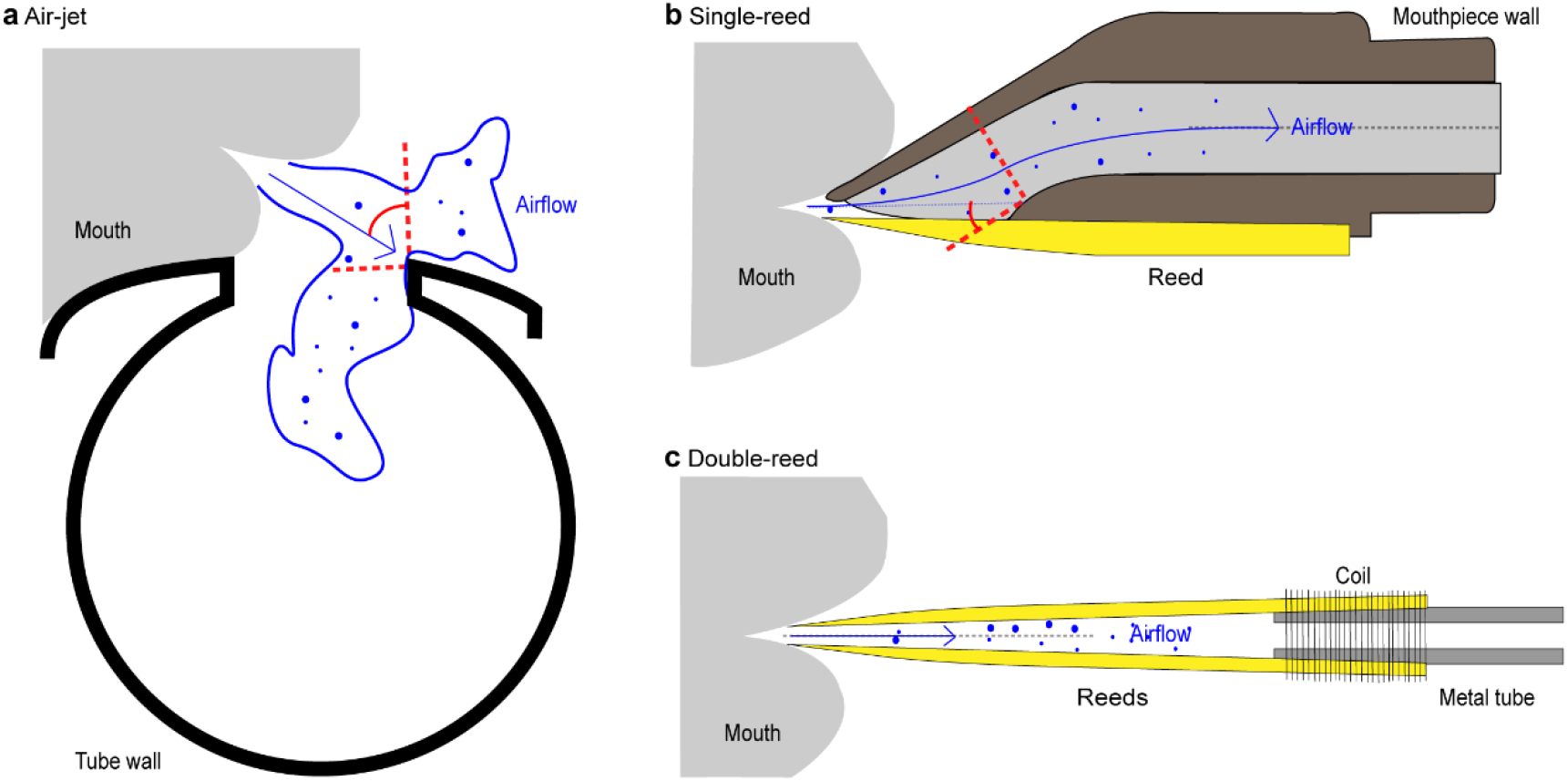
Illustrations of three types of mouthpieces for woodwinds. **(a)** Air-jet mouthpiece used by flute and piccolo. In most situations (Putnik, 1973), the generated aerosols from the mouth directly impinge onto the edge of the embouchure hole with a steep angle (marked by the red dashed lines in the image). This process can lead to a large amount of deposition and air leakage. **(b)** Single-reed mouthpiece used by clarinet and bass clarinet. The aerosols move into the tube through the opening between the edge and reed with a shallower angle in comparison to that in the air-jet woodwinds (marked by the red dashed lines in the image). (**c)** Double-reed mouthpiece used by oboe and bassoon. The generated aerosols from mouth transport into the tube via the opening between two reeds. The airflow is well aligned with these two reeds.

**Fig. S6.**
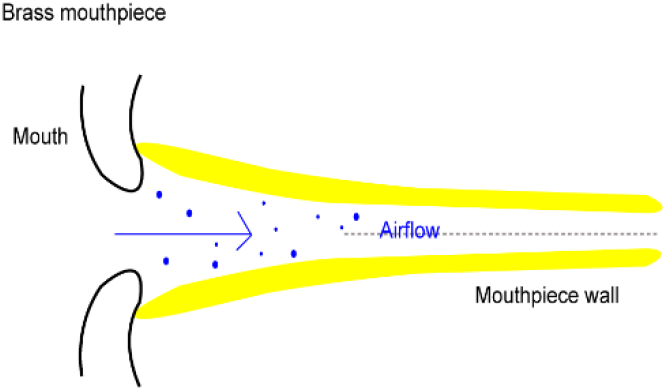
Illustration of mouthpiece for brass. The aerosols generated from the mouth transport straight into the main tube of the brass instrument.

**Fig. S7.**
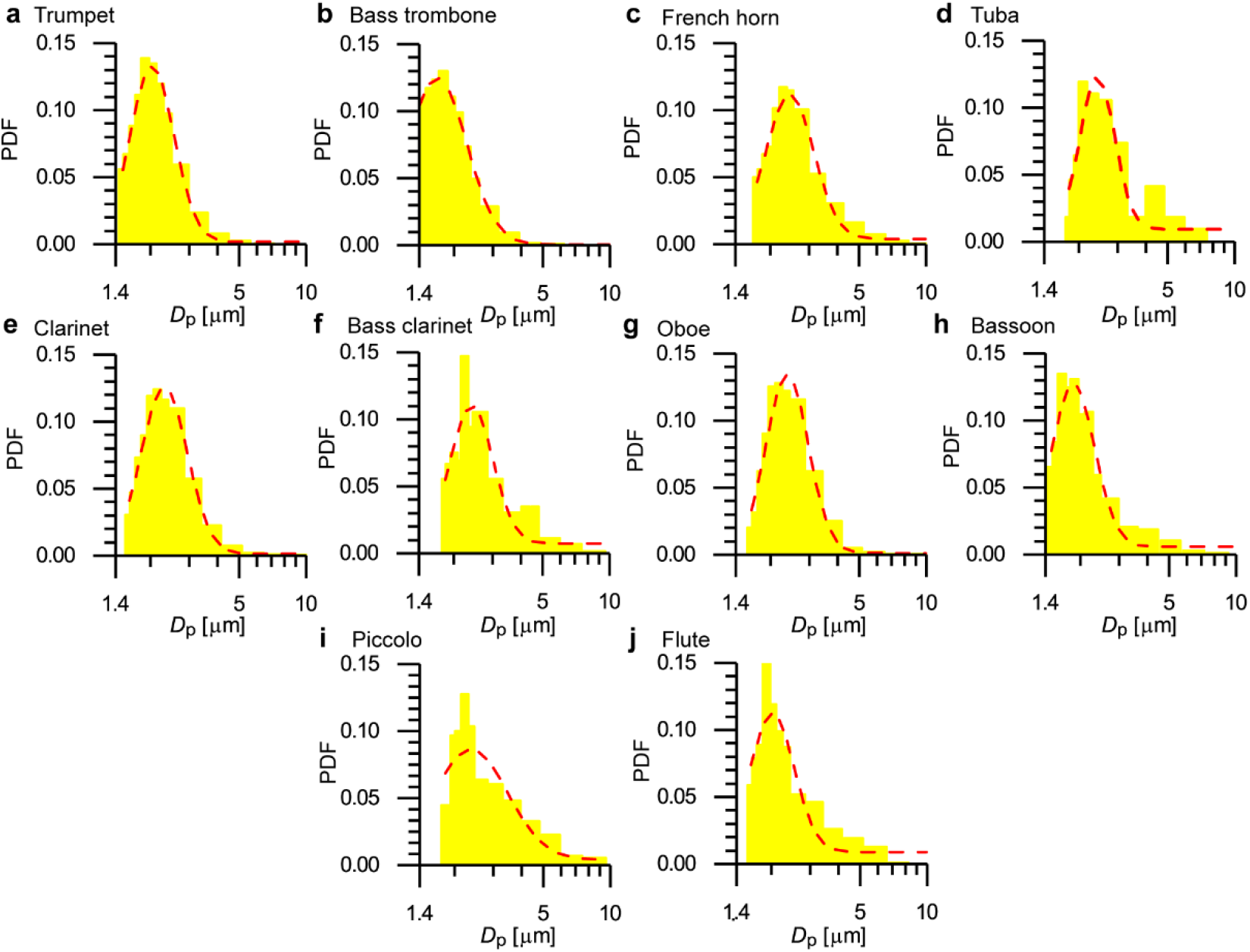
Probability density functions (PDFs) of the aerosol size for each instrument. The red dashed line in each histogram is the log-normal fitting curve of the PDF. In total 16 bins are used in the range from 1.4 to 20 μm. Note that lower bound of 0.5 μm in the raw APS measurement is calibrated to 1.4 μm here based on Eq. (3).

**Fig. S8.**
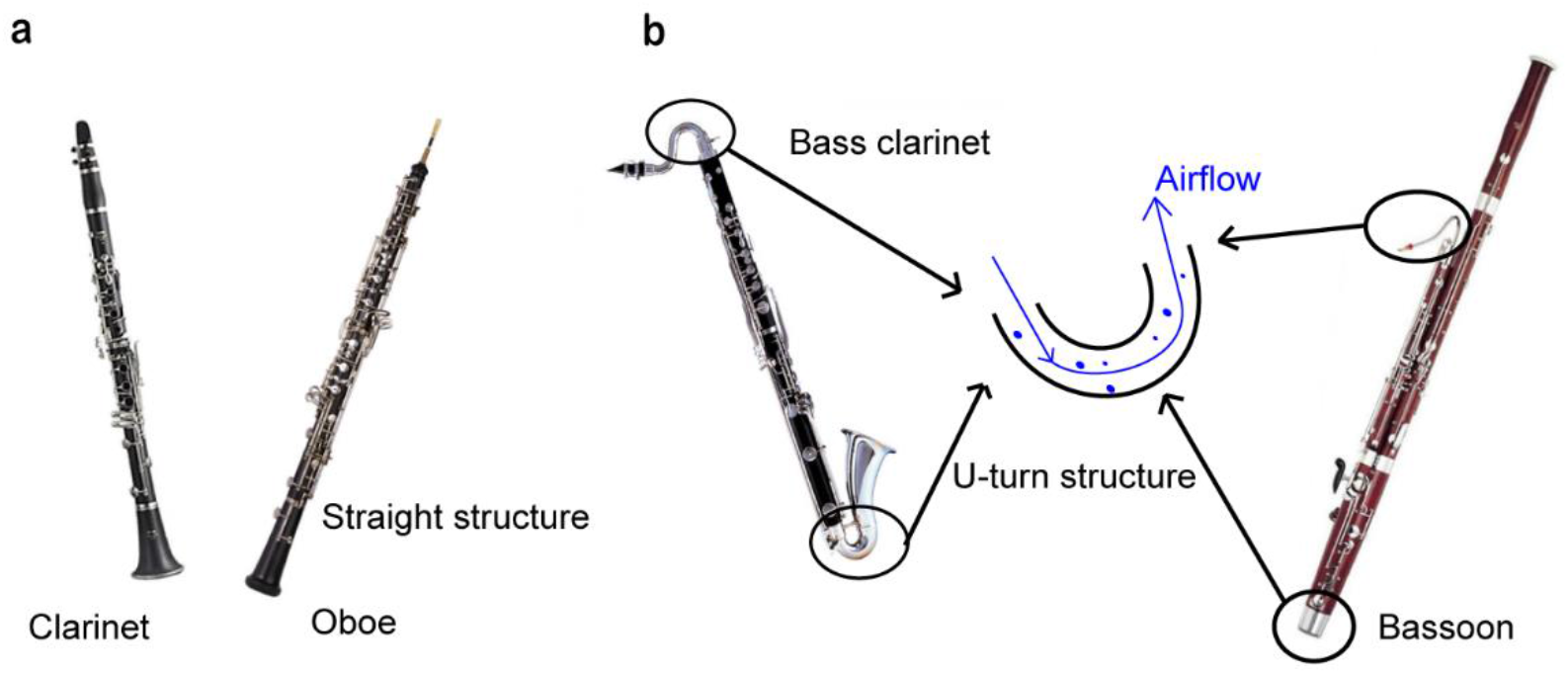
Images of reed instruments. **(a)** Clarinet and oboe. Both clarinet and oboe have straight structures. **(b)** Bass clarinet and bassoon. Bass clarinet has steep turnings near its inlet and outlet while bassoon has a steep turning near the inlet and a U-turning in the middle.

### Supplementary Tables

**Table S1.** A summary of the calibration parameters used in digital inline holography (DIH) for Aerodynamic Particle Sizer (APS) measurements.

| Pressure<br>(psi) | APS |  | DIH |  | DIH/APS |  |  |  |
| --- | --- | --- | --- | --- | --- | --- | --- | --- |
| | $\mu$ | $\sigma$ | $\mu$ | $\sigma$ | mean<br>(ratio) | SD<br>(ratio) | Amplifying<br>ratio (AR) | Shrink<br>ratio (SR) |
| 30 | 0.969 | 0.486 | 1.566 | 0.246 | 1.663 | 0.803 |  |  |
| 35 | 0.933 | 0.466 | 1.513 | 0.247 | 1.653 | 0.841 | 1.66 | 0.84 |
| 40 | 0.808 | 0.463 | 1.397 | 0.252 | 1.670 | 0.872 |  |  |

**Table S2.**
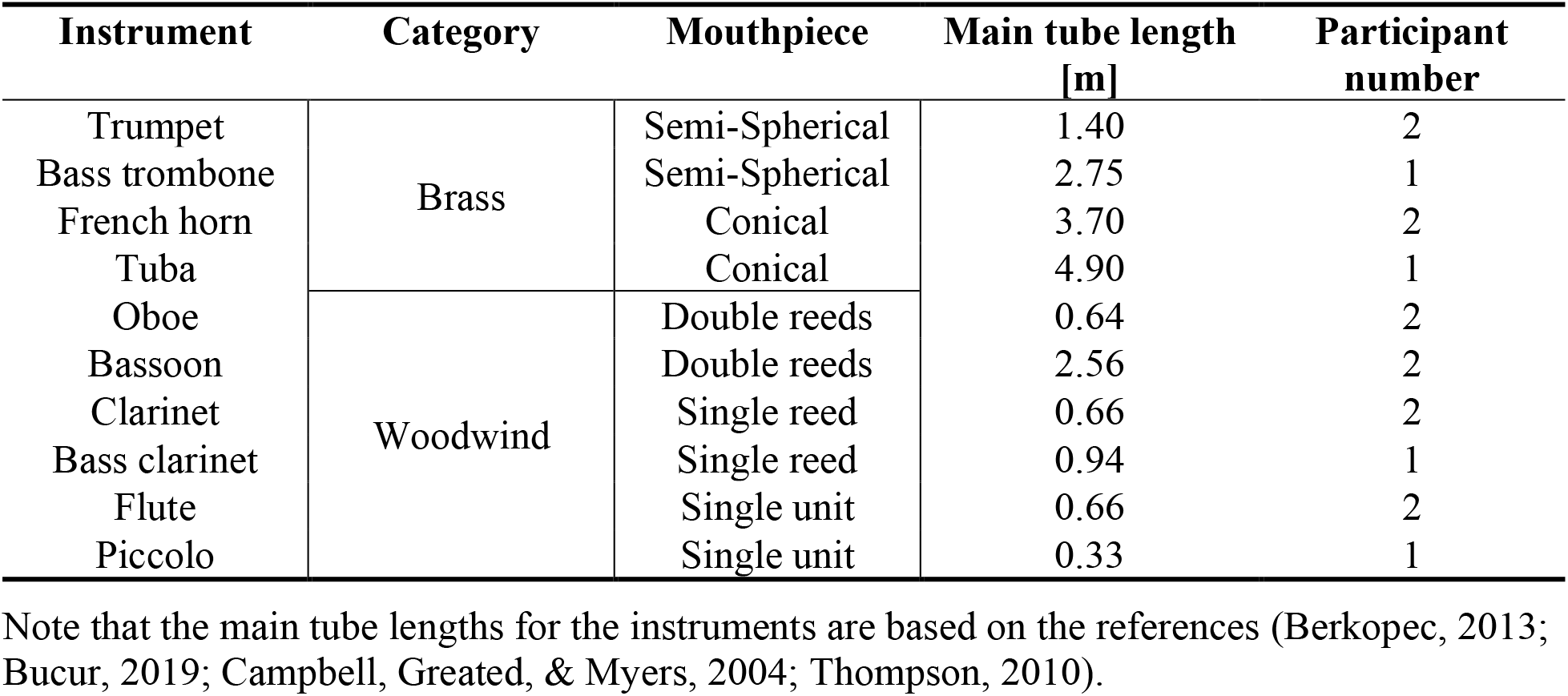
Technical parameters of 10 instruments tested in the study.

| **Instrument** | **Category** | **Mouthpiece** | **Main tube length**<br>**[m]** | **Participant**<br>**number** |
| --- | --- | --- | --- | --- |
| Trumpet | Brass | Semi-Spherical | 1.40 | 2 |
| Bass trombone |  | Semi-Spherical | 2.75 | 1 |
| French horn |  | Conical | 3.70 | 2 |
| Tuba |  | Conical | 4.90 | 1 |
| Oboe | Woodwind | Double reeds | 0.64 | 2 |
| Bassoon |  | Double reeds | 2.56 | 2 |
| Clarinet |  | Single reed | 0.66 | 2 |
| Bass clarinet |  | Single reed | 0.94 | 1 |
| Flute |  | Single unit | 0.66 | 2 |
| Piccolo |  | Single unit | 0.33 | 1 |

Note that the main tube lengths for the instruments are based on the references (Berkopec, 2013; Bucur, 2019; Campbell, Greated, & Myers, 2004; Thompson, 2010).

**Table S3.** Estimation of aerosol deposition time and residence time in the flute and piccolo.

| $D_p$<br>[$\mu\text{m}$] | Flute<br>deposition time[s] | Piccolo<br>deposition time [s] | Flute<br>residence time [s] | Piccolo<br>residence time [s] |
| --- | --- | --- | --- | --- |
| 1.00 | 33.91 | 19.63 | 0.66 | 0.11 |
| 2.00 | 8.48 | 4.91 | 0.66 | 0.11 |
| 3.00 | 3.77 | 2.18 | 0.66 | 0.11 |
| 4.00 | 2.12 | 1.23 | 0.66 | 0.11 |
| 5.00 | 1.36 | 0.79 | 0.66 | 0.11 |
| 6.00 | 0.94 | 0.55 | 0.66 | 0.11 |
| 7.00 | 0.69 | 0.40 | 0.66 | 0.11 |
| 8.00 | 0.53 | 0.31 | 0.66 | 0.11 |
| 9.00 | 0.42 | 0.24 | 0.66 | 0.11 |
| 10.00 | 0.34 | 0.20 | 0.66 | 0.11 |

The residence time (*T*_R_) and deposition time (*T*_D_) of the aerosol in the tube are estimated using Eq. S1 and Eq. S2.

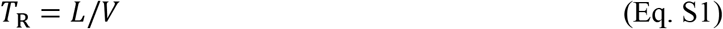

where *L* is the tube length of the instrument, and *V* is the air speed inside the tube. Note that the inlet flowrate for flute and piccolo is assumed to be the same. The air speed inside the flute is set to 1 m/s for the calculation of residence time, which within the range of below 3 m/s (Bamberger, 2002).

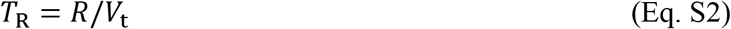

where *R* is the half of the inner diameter of the tube (i.e., half of bore size), and *V*_t_ is the terminal velocity of the aerosol, as given in Eq. S3.

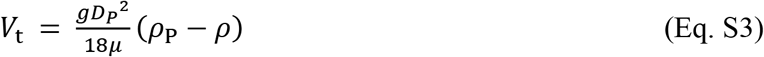

where *μ* is the air viscosity; g is the acceleration due to gravity, *D*_P_ is the aerosol diameter; *ρ*_P_ and *ρ* are the density of aerosols (assuming to be water droplets in this case) and air, respectively. As shown in the Table, for flute, the deposition time of large aerosols (*D*_P_ > 8 μm) is smaller than the corresponding residence time, indicating those large aerosols tend to deposit in the tube before they move out from the instrumental outlet. However, for piccolo, the residence time for the aerosols cross a wide range of sizes is smaller than their deposition time, suggesting that all aerosols can travel out from the outlet. Such trends consolidate the wider size distribution of piccolo than flute in Figs.S7 i and j.

## Notes

### Competing Interest Statement

The authors have declared no competing interest.

### Author Declarations

STUDY00009795: Aerosol generation during human activities was approved by the IRB of the University of Minnesota.

### Summary of Updates

1. add a supplementary figure to show a detailed experimental setup. 2. Section 2.2 updated to provide more detailed experimental setup.

